# A NURSING PHILOSOPHY OF MOTIVATIONAL INTERVIEWING ON TYPE 2 DIABETES MELLITUS: SYSTEMATIC REVIEW

**DOI:** 10.1101/2023.12.19.23300108

**Authors:** Elyk Dwi Mumpuningtias, Moses Glorino Rumambo Pandin, Nursalam Nursalam

## Abstract

**Background:** The management of Type 2 Diabetes Mellitus (T2DM) heavily relies on lifestyle choices and adherence. Opting for a high-risk lifestyle can exacerbate the impact on individuals with T2DM, worsening their condition. These approaches encompass self-care practices, cognitive behavioral strategies, training for family support, and educational initiatives focusing on lifestyle changes. Motivational interviewing is one strategy to changes behavior of T2DM patients.

**Method:** This study reviewed the literature following the PRISMA guidelines (2019– 2023). The exploration involved keyword searches for “motivational interviewing,” “T2DM,” “lifestyle,” and “glycemic index” across databases such as Scopus, Science Direct, and ProQuest. Inclusion criteria encompassed quantitative research design, English language publications, research involving patients with T2DM, motivational interviewing, lifestyle modifications, and glycemic index. This resulted in the identification of 286 papers. A PICO synthesis and grouping method was used to analyze the data, aligned with the Cochrane Handbook for Systematic Reviews. The methodological quality assessment, carried out using the Joanna Briggs Institute Critical Appraisal methodology, indicated that adopting a multidisciplinary approach involving ontological, epistemological, and axiological inquiry is a robust foundation for enhancing health information systems and clinical practice.

**Results:** This study reviewed ten articles focusing on using motivational interviewing techniques to improve T2DM patients so that they can change their lifestyle and manage their glycemic index in the long term. Motivational interviewing can be conducted face to face for an average of 10-12 sessions, approximately3-6 months, both online and through intensive intervention.

**Conclusion:** This study can demonstrate how motivational interviews can improve the behavior of T2DM patients by changing lifestyle and reducing the glycemic index in T2DM patients.

## BACKGROUND

The management of Type 2 Diabetes Mellitus (T2DM) heavily relies on lifestyle choices and adherence. Opting for a high-risk lifestyle can exacerbate the impact on individuals with T2DM, worsening their condition. According to the American Diabetes Association (ADA) guidelines from 2020, successful interventions have the potential to avert the onset of Type 2 diabetes and associated complications. The chronic care model endorsed by the ADA emphasizes patient-centered care, advocating for self-management strategies. This approach involves collaborative planning within a team context, tailored to respect individuals’ unique preferences, values, and specific needs concerning diabetes control (American Diabetes Association, 2020).

Given the prevalence of T2DM and its associated health complications and financial burdens, it becomes crucial to address these challenges indirectly, particularly by encouraging lifestyle modifications. These approaches encompass self-care practices, cognitive behavioral strategies, training for family support, and educational initiatives focusing on lifestyle changes. Implementing effective diabetes management requires lifestyle, behavior, and psychological adjustments. Challenges in adopting behavioral changes can amplify feelings of uncertainty and frustration. Motivational interviewing is an effective collaborative communication style for promoting lifestyle changes in managing T2DM (Bilgin *et al*., 2022).

Motivational interviewing (MI) is one strategy to enhance individuals’ lifestyle choices and adherence to T2DM. MI aids individuals in addressing uncertainties surrounding health and illness linked to their behavior. It is an approach that facilitates adherence to treating chronic conditions like diabetes mellitus (DM). Health professionals utilize MI effectively to enhance an individual’s motivation for continuous treatment and care, thereby boosting compliance. Numerous studies demonstrate the positive impact of MI on individual management, showing its assistance in regulating metabolic variables. Systematic reviews and meta-analyses in the literature indicate that MI contributes to reducing HbA1c values, a critical parameter in diabetes management, and positively influences an individual’s dietary behavior (Ramadan *et al*., 2019; Celano *et al*., 2019).

## METHOD

### Inclusion and axclusion criteria

The methodology involves a literature review exploring pertinent peer-reviewed articles available in Scopus, ProQuest, and ScienceDirect. The inclusion and exclusion criteria are applied by the search and selection criteria. (1) Using PICOS (population, intervention, comparison, outcome, and study design) is one of the inclusion criteria. Population (P) denotes research involving T2DM patients. Intervention (I) encompasses all studies on motivational interviewing in T2DM patients. Motivational interviewing on T2DM patients is crucial to improving lifestyle and adherence and lowering glycemic index. Comparison (C) requires a comparative group for motivational interviewing on T2DM patients. Outcome (O) necessitates research containing analysis supporting motivational interviewing on T2DM patients.

All forms of quantitative studies fall under the category of research design (S). (2) Publications within the past five years, from 2019 to 2023, are considered for inclusion. (3) All publications included must be in English. Exclusion criteria involve protocol studies, conference presentations, editorial and review papers, case reports, case series, and applied design and development.

### Search strategy

The research methodology employed in this study involves a literature review. The review article delves into the philosophical examination of applying motivational interviewing to individuals with T2DM, exploring aspects related to their lifestyle and glycemic index from epistemological and ontological perspectives. The researchers utilized the Systematic Review and Meta-Analysis Preferred Reporting Items (PRISMA) guidelines to structure the survey (Page *et al*., 2021). The literature search strategy encompassed relevant keywords such as “motivational interviewing,” “T2DM,” “patients,” “lifestyle,” and “glycemic index,” with articles sourced from databases including Scopus, Science Direct, and ProQuest, covering publications from 2019 to 2023. The primary objective of this literature analysis method was to investigate, assess, and amalgamate insights from pertinent literature, aiming to gain deeper insights into how motivational interviewing influences the lifestyle and glycemic index of individuals diagnosed with T2DM within the philosophical framework of nursing science. The study identified 286 articles from databases, with Science Direct contributing 231 articles, ProQuest providing 15, and Scopus contributing 40. The literature search adhered to the minimum requirement of combining the top three databases, as suggested in systematic survey practices (Rethlefsen *et al*., 2021). Key search terms were selected to align with the research topic, employing keywords like “motivational interviewing,” “T2DM,” “patient,” “lifestyle,” and “glycemic index,” combined with the boolean operators “AND/OR.”

### Selction of study

All authors (EDM, NN, and MG) searched academic databases comprehensively. Following this, we proceeded with the article selection process. Reviewer (EDM) utilized Rayyan, an intelligent tool for systematic reviews, to select publications. Rayyan, a mobile and online application designed for systematic reviews (Ouzzani *et al*., 2016), has demonstrated efficacy in streamlining the review process and easing the workload for reviewers (J. Li *et al*., 2023). Articles filtered from three extensive databases were imported into the Rayyan platform, resulting in an initial identification of 286 articles. Subsequently, duplicate articles were removed through a meticulous examination. The author (EDM) then individually assessed the titles and abstracts of the remaining articles, adhering to predetermined inclusion criteria. Specifically, the focus was on titles and abstracts related to motivational interviewing on T2DM patients. This rigorous screening process identified ten relevant studies from the significant databases. These selected studies were independently read in their entirety, and upon meeting the criteria, they were included in this systematic review (Figure 1).

**Figure 1.**
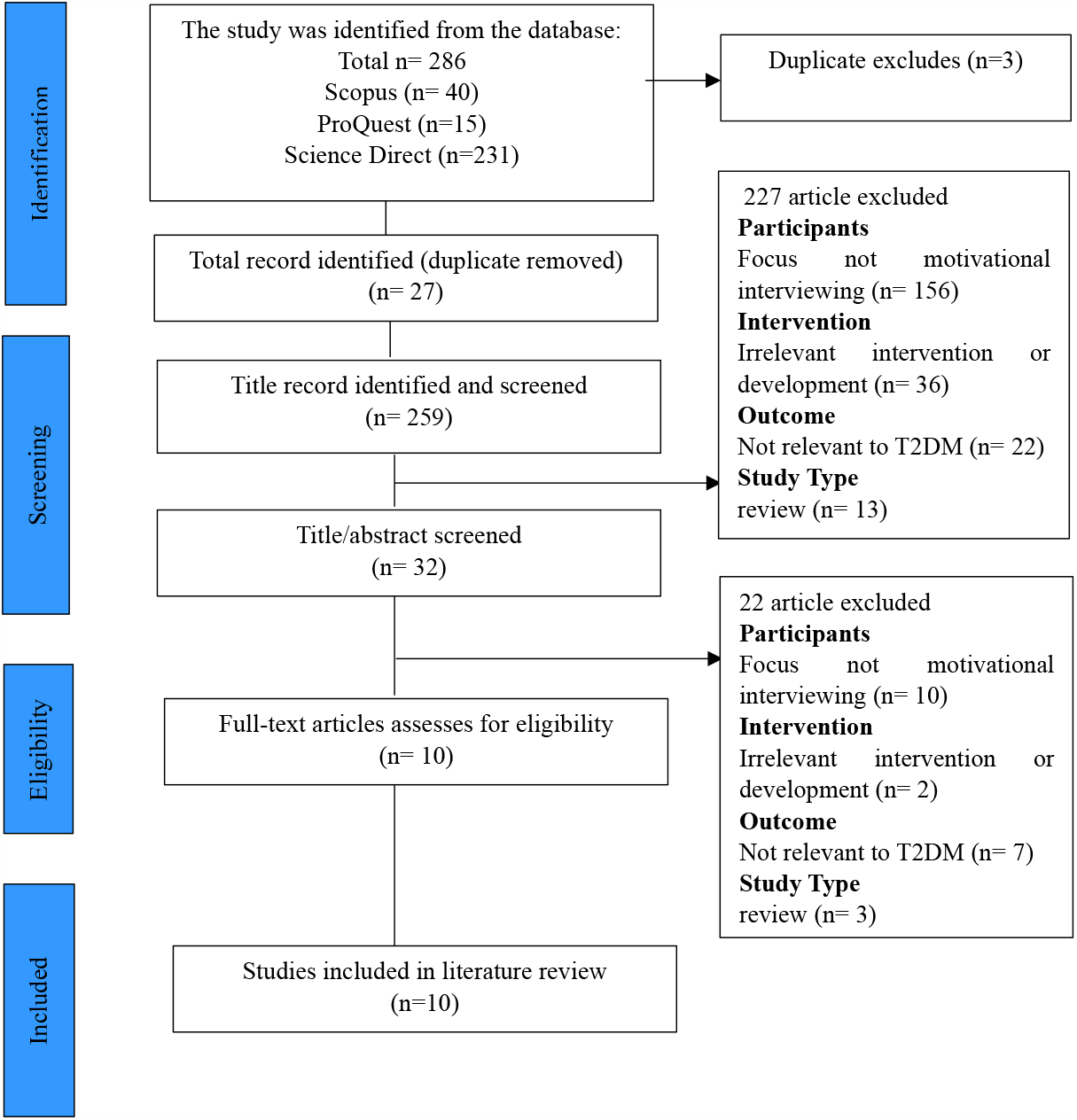
Process flow diagram for PRISMA literature search and filtering.

**Table 1.**
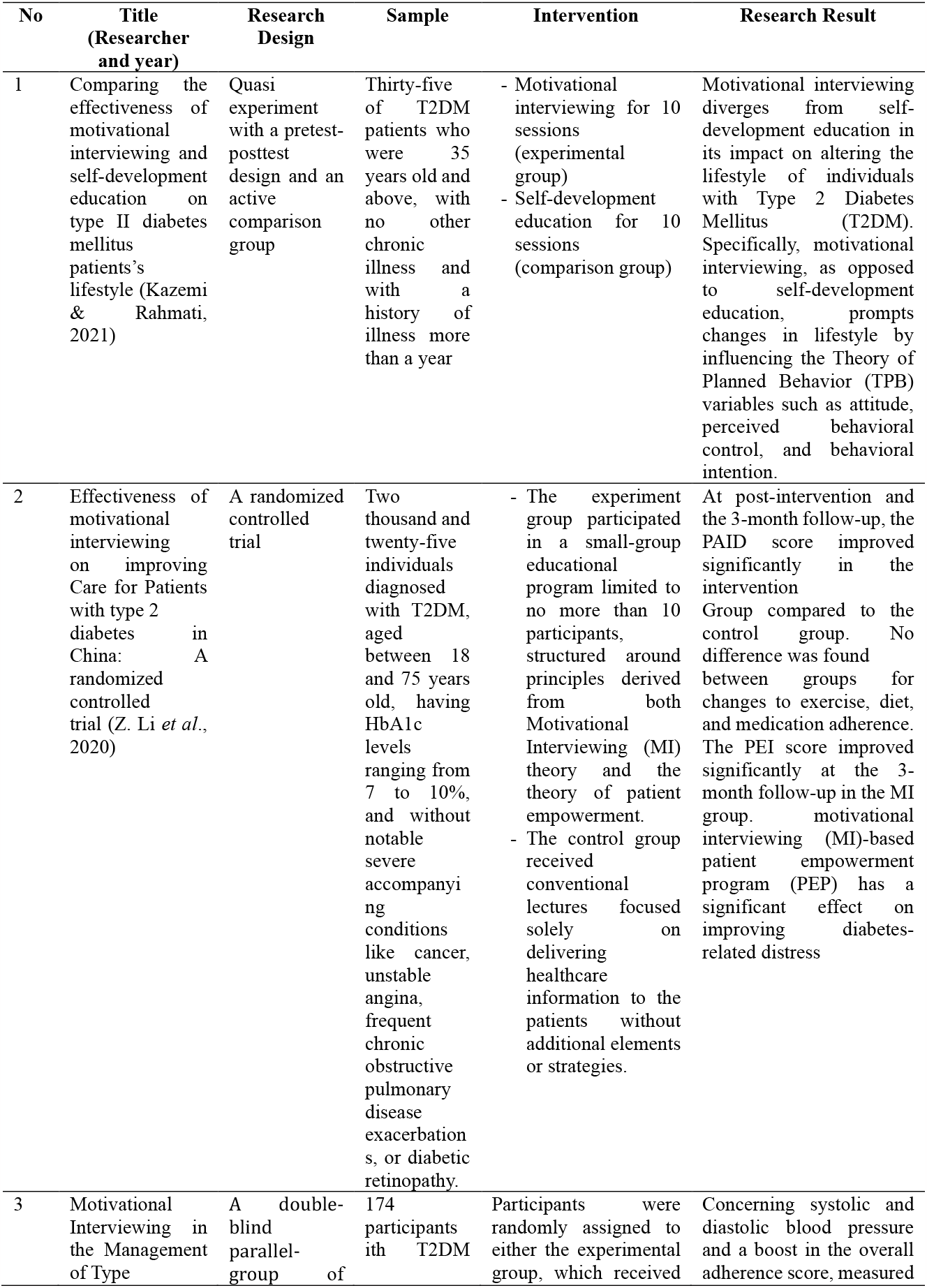
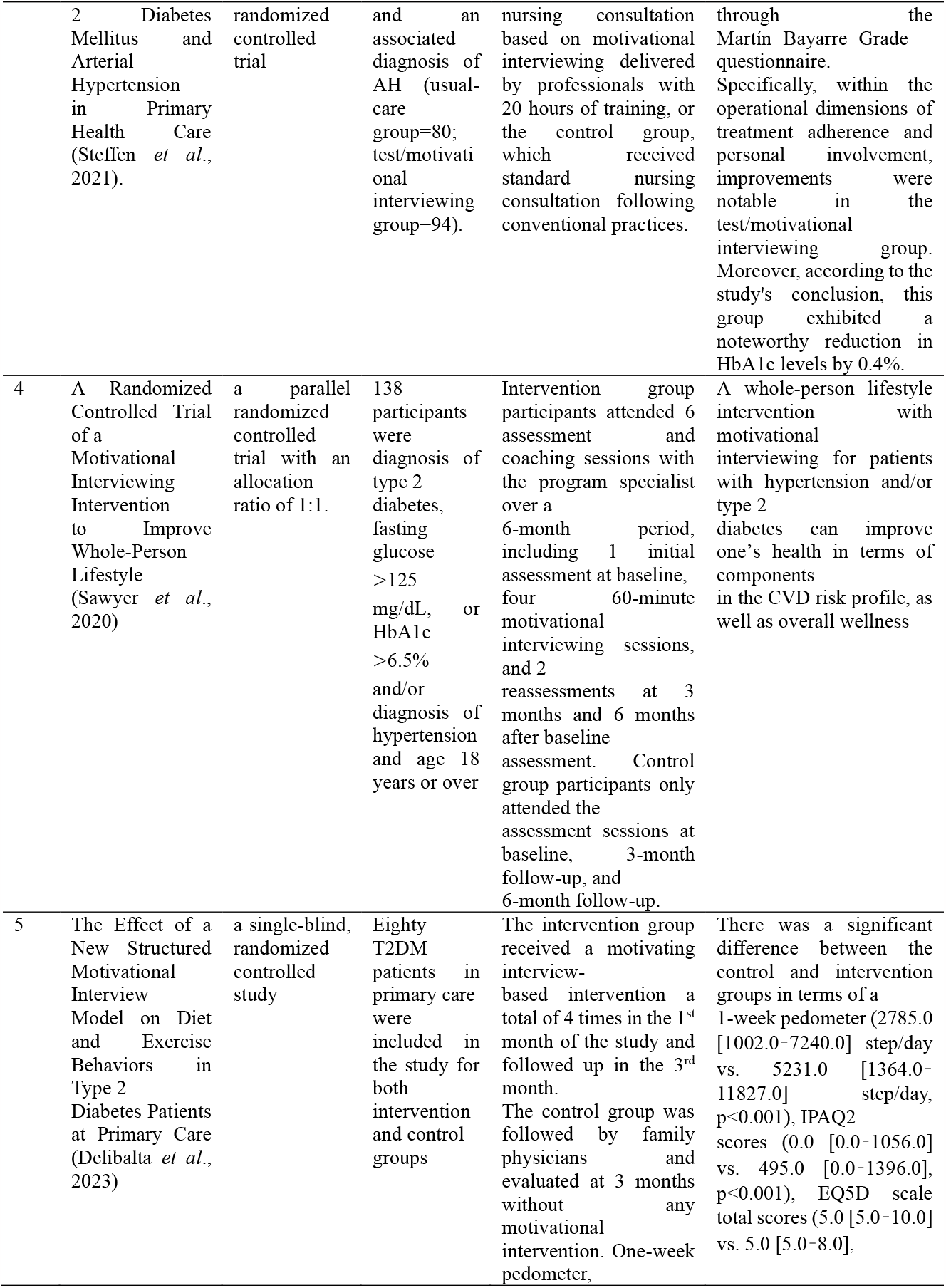
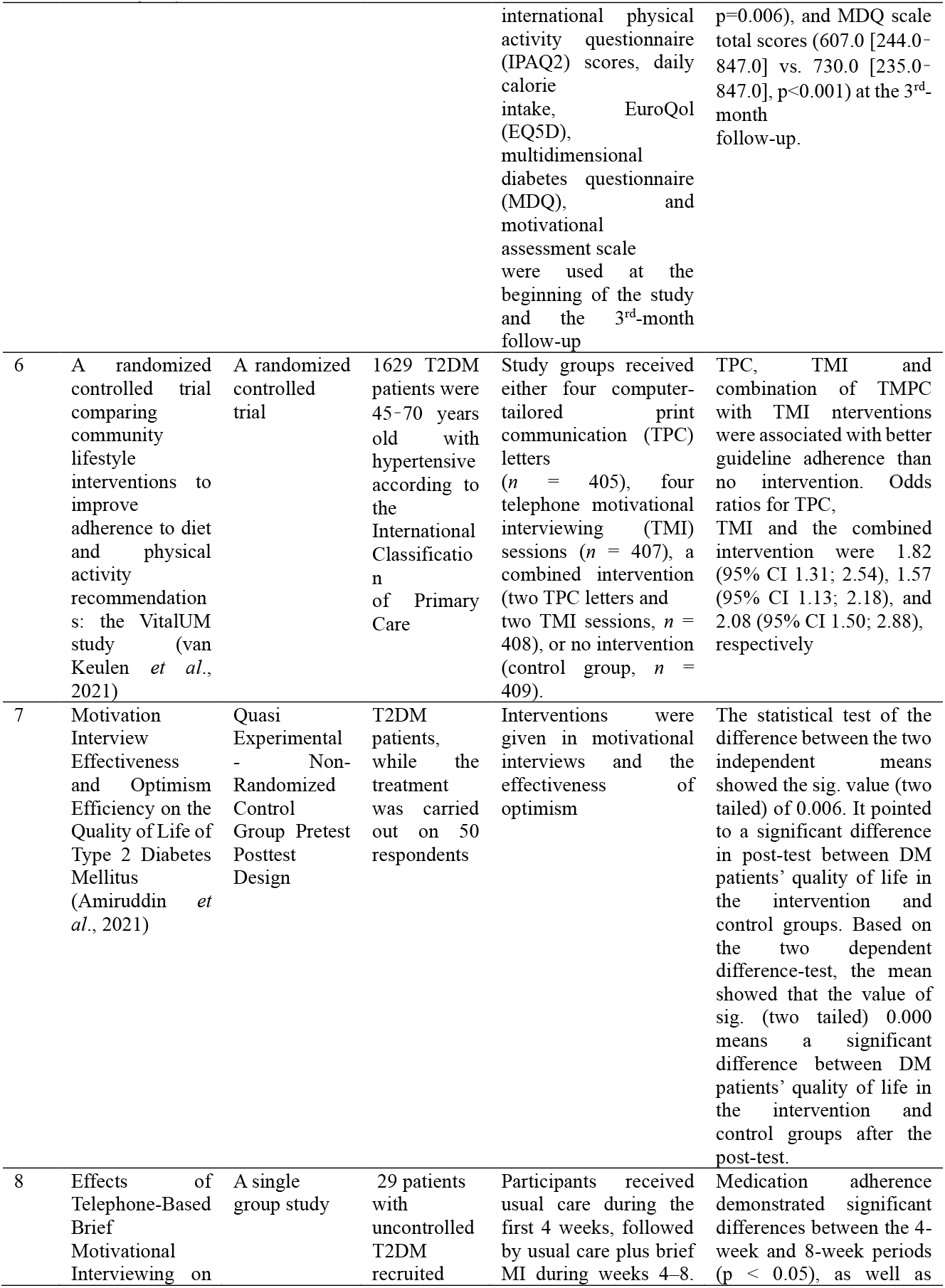
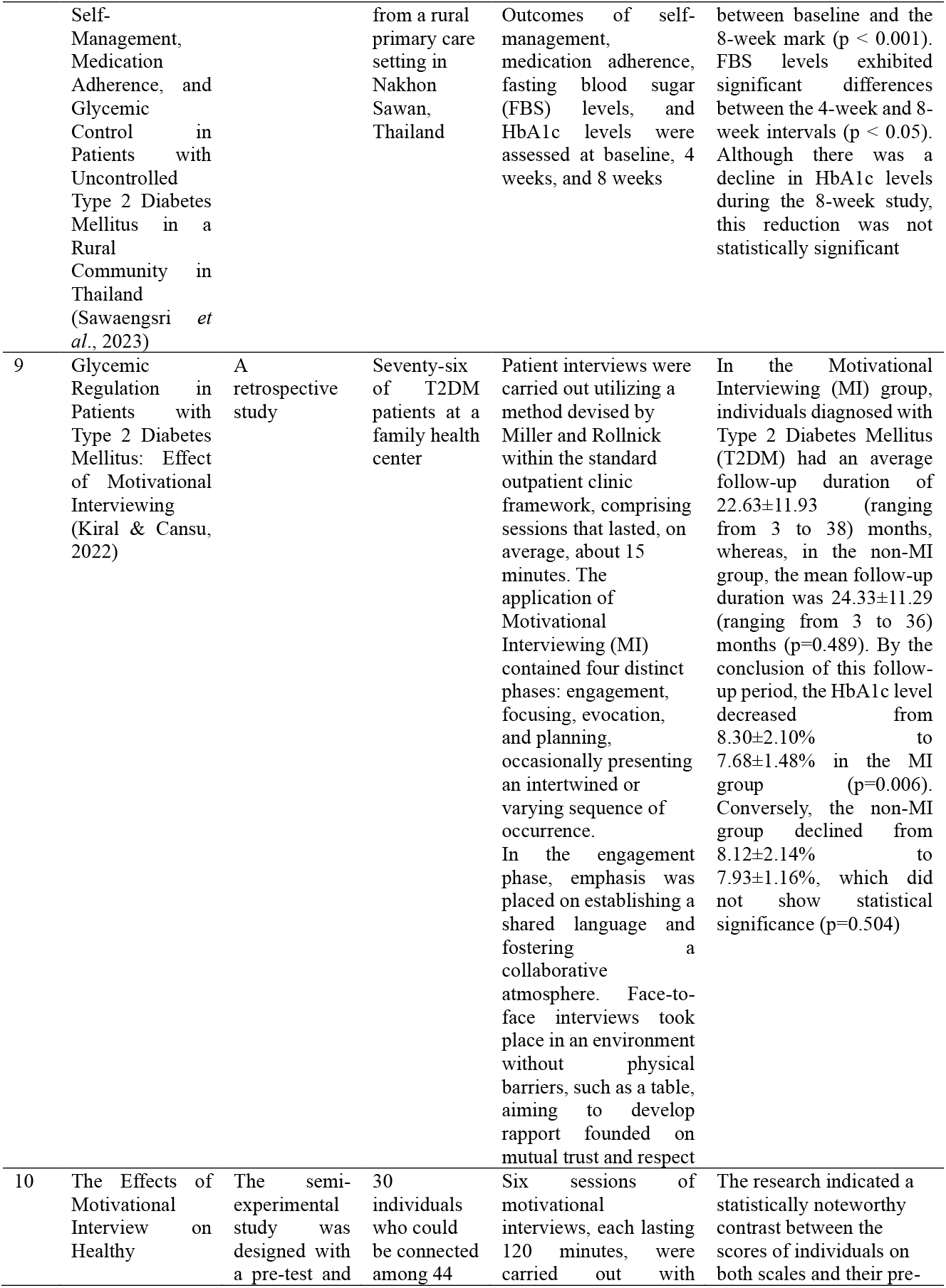
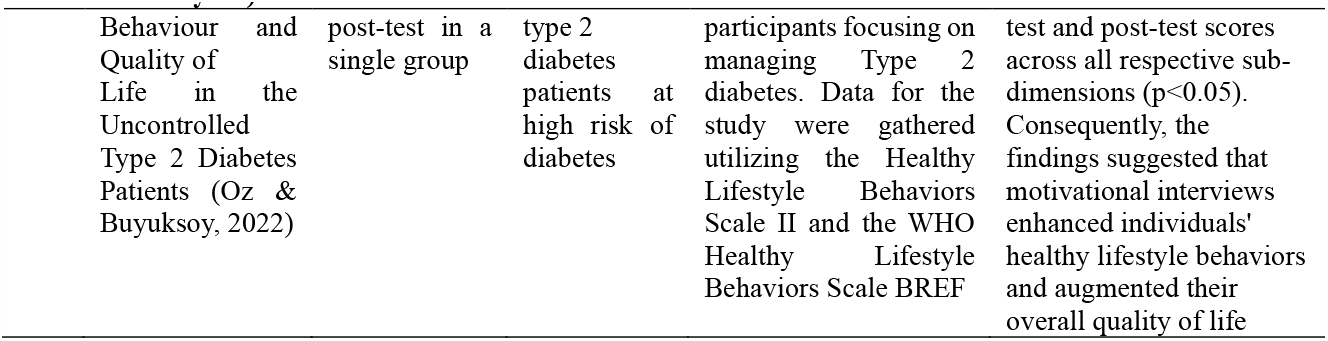
Characteristics of reviewed studies.

### Risk of bias and research quality

The analysis adopts PICO synthesis as its methodology. The initial stage of this synthesis involves extracting the study characteristics—such as population, intervention, comparison, results, and research design—from each acquired article. Subsequently, the Cochrane Handbook for Systematic Reviews (Cumpston *et al*., 2021) facilitates categorizing these features. The authors then assess the research quality by examining the risk of bias. This assessment aims to evaluate the methodological rigor of the studies and determine the extent to which each study has mitigated potential biases in its design, execution, and analysis. Systematic reviews employ the Joanna Briggs Institute (JBI) critical assessment tool to gauge the risk of bias, aligning it with the specific research design used in each study (Munn *et al.*, 2020). The researchers also used the search criteria of this review literature with the PRISMA flow diagram to obtain a suitable and eligible research article.

## RESULTS AND DISCUSSION

The researchers used the PRISMA flow diagram and literature review search criteria to identify appropriate research articles that met the requirements. These articles aim to understand the effectiveness of motivational interviewing in improving lifestyle, compliance, and reducing the glycemic index of T2DM patients. This approach further strengthens the nursing science philosophy’s importance in addressing these needs.

### Ontological study of motivational interviewing on improving lifestyle and lowering glycemic index of T2DM patients

The structure and hierarchy of concepts in motivational interviewing are analyzed in an ontological study of patients with T2DM. As a branch of philosophy, ontology examines the truth of the existence and relationships between entities. In ontological studies from motivational interviews of T2DM patients, the focus is on lifestyle changes, including self-care techniques, cognitive behavioral techniques, family support training, and lifestyle education (Bakır *et al*., 2021)

This research provides a conceptual basis for the design application of motivational interviewing to T2DM patients from an ontological perspective. By understanding its ontological structure, health professionals can effectively integrate these concepts into clinical practice, improving the quality of patient care for T2DM patients and supporting further development of efficient health information systems. Improving the quality of care for T2DM patients involves cross-sectoral collaboration, including medical personnel, patients, families, and health service facilities. This includes routine training of medical personnel, formation of a holistic care team, the implementation of standard care protocols, periodic evaluation and monitoring, integration health technology, education for patients and families, and coordinated rehabilitation programs. As revealed in the research, motivational interviewing is defined as finding and eliminating doubts and increasing motivation to change. Motivational interviewing includes collaborative efforts, client-centeredness, non-judgment, building trust, reducing resistance, increasing readiness to change, increasing self-efficacy, increasing perceptions of incongruity, reflective listening, conversational invitations to change, discovery, and empathetic listening (Oz & Buyuksoy, 2022; (El-Sappagh *et al*., 2018).

### Epistemological study of motivational interviewing on improving lifestyle and lowering glycemic index of T2DM patients

Epistemological research concerning motivational interviewing in patients with Type 2 Diabetes Mellitus (T2DM) aims to delve deeply into the nature of knowledge associated with this approach. Epistemology, a philosophical discipline, examines the sources, boundaries, and fundamental learning principles. Within the context of epistemological investigation concerning motivational interviewing in T2DM patients, the emphasis is placed on how knowledge pertinent to this process is acquired, comprehended, and justified. Through an epistemological lens, this research provides insight into the origins and attributes of knowledge concerning motivational interviewing in T2DM patients, explicitly aiming to enhance lifestyle improvements and glycemic control. This approach seeks to improve comprehension of how this knowledge is applied and assessed in healthcare practices, thereby stimulating considerations on improving these processes using an epistemological method in the exploration and study of motivational interviewing in T2DM patients (Adailton da Silva *et al*., 2018).

To acquire, comprehend, and authenticate knowledge associated with applying motivational interviewing to enhance lifestyle and glycemic control in T2DM patients, we can undertake several steps that can be conducted by engaging in an in-depth review of the latest health literature, including books, journals, and research articles discussing the application of motivational interviewing in T2DM patients. This step aids in understanding current scientific foundations and practices in this domain. Firstly, participate in seminars, conferences, or workshops on T2DM management to engage with experts directly, interact, and exchange experiences with fellow healthcare professionals. In addition, seek guidance from healthcare practitioners such as doctors, nurses, psychologists, and other professionals experienced in treating T2DM patients to gain practical insights and learn from their clinical expertise. Furthermore, utilize reliable online resources like health organization websites, medical institutions, or research centers offering up-to-date and verified information for managing T2DM patients. In conclusion, refer to guidelines and service standards issued by national and international health institutions, like the American Diabetes Association (ADA), to align with recognized protocols and practices (Mureyi *et al*., 2022).

### Axiological of motivational interviewing on improving lifestyle and lowering glycemic index of T2DM patients

An axiological exploration concerning the application of motivational interviewing to enhance lifestyle and glycemic control in T2DM patients will involve an analysis of the moral values and principles intrinsic to the motivational interviewing process in this patient population. Axiology, a philosophical branch, specifically investigates moral values. The axiological scrutiny involving motivational interviewing in T2DM patients focuses on understanding how these values impact and shape nursing practices. The aim is to pinpoint the fundamental values that underpin motivational interviewing in T2DM patients, namely Acceptance, Partnership, Compassion, and Evocation. The analysis aims to elucidate how these values influence interactions between nurses and patients throughout the therapeutic process, particularly their impact on communication, collaborative decision-making, and empowering behavioral changes (Van Wormer, 2007).

According to research, education is of significant value for T2DM patients due to its role as an indicator of patients’ comprehension of care, self-management, and glycemic control. The willingness of T2DM patients to embrace health information correlates with their adoption of healthier lifestyle behaviors, leading to an enhanced quality of life. Maintaining controlled blood glucose levels and stable health status aligns with patients’ openness to health education. Besides education, cultural and personal values significantly influence motivational interviewing’s efficacy in altering individuals’ behaviors. Subjective value judgments and the prevailing societal culture are pivotal in shaping individuals’ daily practices (Steffen *et al*., 2021). Studies have indicated that cultural aspects are critical considerations for the effectiveness of motivational interviews, particularly in influencing behavior change. For instance, research illustrated that motivational interviews weren’t effective in promoting healthy lifestyle behaviors among individuals at risk of diabetes in South Asia. Moreover, most individuals did not continue with the discussions, indicating the necessity to account for cultural characteristics to ensure the effectiveness of interventions (Magill & Hallgren, 2019; Amiruddin *et al*., 2021).

### Motivational interviewing on improving lifestyle and lowering glycemic index of T2DM patients

Lifestyle is crucial in managing Type 2 Diabetes Mellitus (T2DM). Opting for a high-risk lifestyle can exacerbate its impact on individuals with T2DM. Implementing self-care techniques, cognitive behavioral strategies, family support training, and lifestyle education becomes imperative to enhance the lifestyle and glycemic control of T2DM patients. Motivational interviewing is an alternative approach to foster behavioral changes among individuals with T2DM. This technique identifies and resolves uncertainties while amplifying the motivation to instigate change. Components integral to motivational interviewing encompass collaborative efforts, client-centeredness, non-judgmental attitudes, building trust, mitigating resistance, enhancing readiness for change, bolstering self-efficacy, recognizing discrepancies, engaging in reflective listening, employing conversational prompts for change, fostering discovery, and empathetic listening (Delibalta *et al*., 2023). Regarding behavioral alterations correlated with the lifestyle of T2DM patients, motivation proves to be more effective than knowledge. Among the five factors influencing behavior—awareness, social norms, perceived behavioral control, attitudes, and behavioral intentions—motivation outweighs the ability to facilitate changes to a patient’s lifestyle. While increased knowledge contributes to positive lifestyle behaviors, particularly in chronic conditions like diabetes, it’s more impactful in endorsing positive behaviors than eliminating negative habits linked to adverse physical conditions. Overcoming ingrained negative habits and burdensome lifestyles is significantly more influenced by motivation than mere knowledge (Kazemi & Rahmati, 2021).

Motivational interviewing is a practical collaborative communication approach to foster lifestyle modifications in Type 2 Diabetes Mellitus (T2DM) management. The research indicated noticeable differences between the control and test/MI groups in blood pressure and compliance scores following an average six-month follow-up. Notably, there was a significant reduction of 0.4% in HbA1c levels among participants who underwent motivational interviewing. This decrease holds clinical significance, particularly considering motivational interviewing is integrated as a supplementary and routine component in diabetes management within primary healthcare settings. In primary healthcare, nursing consultation based on motivational interviewing was proven to be a more efficacious care strategy than standard care in enhancing blood pressure levels and compliance rates among individuals diagnosed with T2DM (Steffen *et al*., 2021).

## CONCLUSION

Review of the literature with a focus on improving lifestyle and lowering glycemic index of T2DM patients by motivational interviewing. This study can demonstrate how motivational interviews can improve the behavior of T2DM patients by changing lifestyle and reducing the glycemic index in T2DM patients.

## Data Availability

All data produced in the present work are contained in the manuscript

## ACKNOWLEDGMENTS

Thank you to the nursing lecturers at Airlangga University and very grateful that the Faculty of Nursing at Airlangga University has provided facilities for this research

## CONFLICT OF INTEREST

None

## Notes

### Competing Interest Statement

The authors have declared no competing interest.

### Funding Statement

This study did not receive any funding

